# Identifying Sepsis Subphenotypes via Time-Aware Multi-Modal Auto-Encoder

**DOI:** 10.1101/2020.07.26.20162214

**Authors:** Changchang Yin, Ruoqi Liu, Dongdong Zhang, Ping Zhang

## Abstract

Sepsis is a heterogeneous clinical syndrome that is the leading cause of mortality in hospital intensive care units (ICUs). Identification of sepsis subphenotypes may allow for more precise treatments and lead to more targeted clinical interventions. Recently, sepsis subtyping on electronic health records (EHRs) has attracted interest from healthcare researchers. However, most sepsis subtyping studies ignore the temporality of EHR data and suffer from missing values. In this paper, we propose a new sepsis subtyping framework to address the two issues. Our subtyping framework consists of a novel Time-Aware Multi-modal auto-Encoder (TAME) model which introduces time-aware attention mechanism and incorporates multi-modal inputs (e.g., demographics, diagnoses, medications, lab tests and vital signs) to impute missing values, a dynamic time wrapping (DTW) method to measure patients’ temporal similarity based on the imputed EHR data, and a weighted k-means algorithm to cluster patients. Comprehensive experiments on real-world datasets show TAME outperforms the baselines on imputation accuracy. After analyzing TAME-imputed EHR data, we identify four novel subphenotypes of sepsis patients, paving the way for improved personalization of sepsis management.

**CCS CONCEPTS:** •**Applied computing** → **Health informatics**; • **Social and professional topics** → *Medical records*; • **Mathematics of computing** → Time series analysis.

**ACM Reference Format:** Changchang Yin, Ruoqi Liu, Dongdong Zhang, and Ping Zhang. 2020. Identifying Sepsis Subphenotypes via Time-Aware Multi-Modal Auto-Encoder.

## 1 INTRODUCTION

Sepsis, defined as life-threatening organ dysfunction in response to infection, contributes to up to half of all hospital deaths and is associated with more than $24 billion in annual costs in the United States [11]. Treating a septic patient is highly challenging because individual patients respond differently to medical interventions. Identification of sepsis subphenotypes may lead to more precise treatments and more targeted clinical interventions.

Over the past few decades, the rapid growth in volume and diversity of electronic health records (EHRs) makes it possible to apply machine learning and data mining methods to subtype patients based on their EHR data. EHRs are temporal sequence data and consist of demographics, diagnoses, medications, lab results, vital signs, and other information, as is shown in Figure 1. Existing sepsis subtyping models [8, 14] cluster patients based on the aggregations of important clinical variables (e.g., heart rate and respiratory rate) during the first day in ICU stays. There are two main limitations of the existing studies. (i) The existing sepsis subtyping frameworks [8, 14] adopt the aggregation of clinical variables to compute the patient similarity, which ignores the variables’ temporality, an important characteristic of EHR data. (ii) Most existing subtyping models [8, 14, 20, 23] suffer from missing values and some models [8] even exclude the patients with variables with high missing rates. However, the variables used to subtype sepsis patients have various missing rates. Especially in the early stage of patient admissions and ICU stays, many variables’ missing rates are relatively high. Both the exclusion of patients with high missing rate data and simple imputation models (e.g., mean imputation used in [20] and MICE [3] used in [14, 23]) are not very suitable for sepsis subtyping.

**Figure 1:**
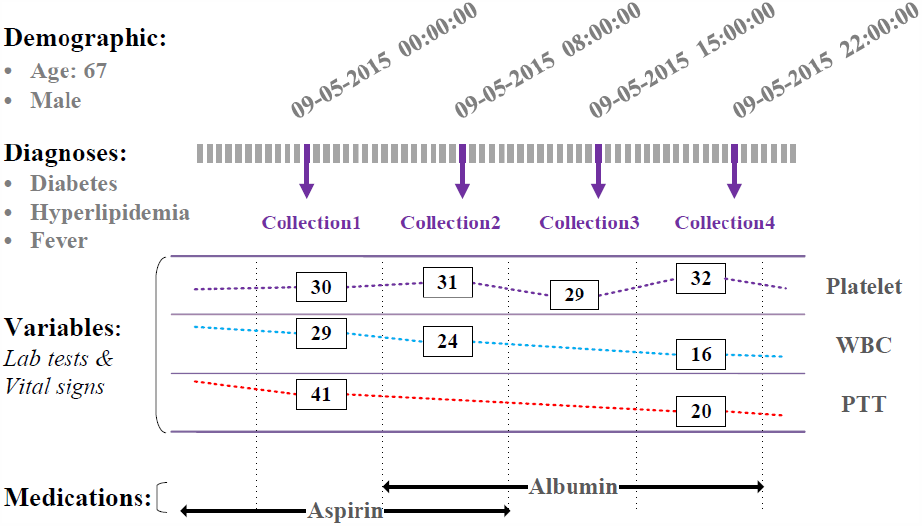
An example segment of a patient’ EHR data inside an admission. The patient has demographics information, a set of diagnoses, lots of collections of variables (including lab tests and vital signs), a set of medications. In each collection, different variables may have missing values. Time spans between two successive collections can vary. The medications are prescribed at different time, and the prescription periods can also vary. Such time irregularity and missing values result in a significant challenge in sepsis subtyping from EHRs.

In this study, we propose a novel sepsis subtyping framework to address the issues. Our subtyping framework consists of three steps to group sepsis patients. The first step is to impute missing values with a novel Time-Aware Multi-modal auto-Encoder (TAME) model. TAME encodes multi-modal inputs (e.g., demographics, diagnoses, medications, lab tests, and vital signs) and decodes the values of sepsis-related variables. We propose a time gap embedding and time-aware attention for TAME to take account of the irregular time gaps between collections and variables’ longitudinal information. Moreover, to handle various numbers of observed values in different collections and combine multi-modal data, we propose a new value embedding to project variables and their values into an embedding space while retaining the values’ continuity so that similar values have similar embeddings. The second step is to adopt dynamic time wrapping (DTW) [13] to calculate the temporal similarity between patients with the imputed data. The third step is to cluster the patients with weighted k-means, which assigns weights for the patients in a group when computing the distances between the group and patients.

To demonstrate the efficacy of the proposed model, we conduct imputation experiments on two publicly available datasets: DACMI^1^ and MIMIC-III [9]. The results show our model outperforms the baselines. Moreover, based on the imputed EHR data, we group sepsis patients using their first 24 hours’ worth of data in ICUs into four meaningful subphenotypes. The experimental results show that both the well-imputed EHR data and the weighted k-means algorithm can significantly improve the subtyping performance. Finally, we analyze the characteristics of the four subphenotypes and discuss their potential for sepsis personalized medicine.

In sum, our contributions are as follows:

- We design a new patient subtyping framework that integrates clinical data imputation model TAME, temporal similarity analysis with DTW, and a weighted k-means method to identify sepsis subphenotypes on EHR data.
- We develop a new imputation model TAME that can handle multi-modal inputs and incorporate cross-modal relations.
- We incorporate value embedding to represent each variable value into a vector so that TAME can handle varying numbers of missing values across collections.
- We introduce time embedding and time-aware attention to TAME to consider collections’ irregular time intervals and variables’ longitudinal information.
- Finally, we demonstrate the effectiveness of our methods experimentally on two real-world EHR data. By using only EHR data from the first 24 hours of patients’ ICU stay, we identify four novel subphenotypes with different clinical characteristics and mortality trajectories, paving the way for personalized medicine for sepsis.

The rest of the paper is organized as follows. In Section 2, we describe technical details of the proposed sepsis subtyping framework. In Section 3, we conduct experiments on two real-world EHR datasets. We review the related studies in Section 4. Section 5 concludes our work.

## 2 METHODOLOGY

In this section, we propose a new time-Aware Multi-modal auto-Encoder (TAME) model to impute missing values in EHR data. Then we leverage Dynamic Time Wrapping (DTW) [13] to compute patient similarity. Finally, we present a weighted k-means to identify subgroups of sepsis patients.

### 2.1 Data Imputation with TAME

TAME takes multi-modal data as inputs by embedding them into a same space. Then a max-pooling layer is used to combine the multi-modal data’s information and output fixed-size vectors, which are sent to BiLSTM to model the time series data and predict the missing values. The framework of TAME is shown in Figure 2.

**Figure 2:**
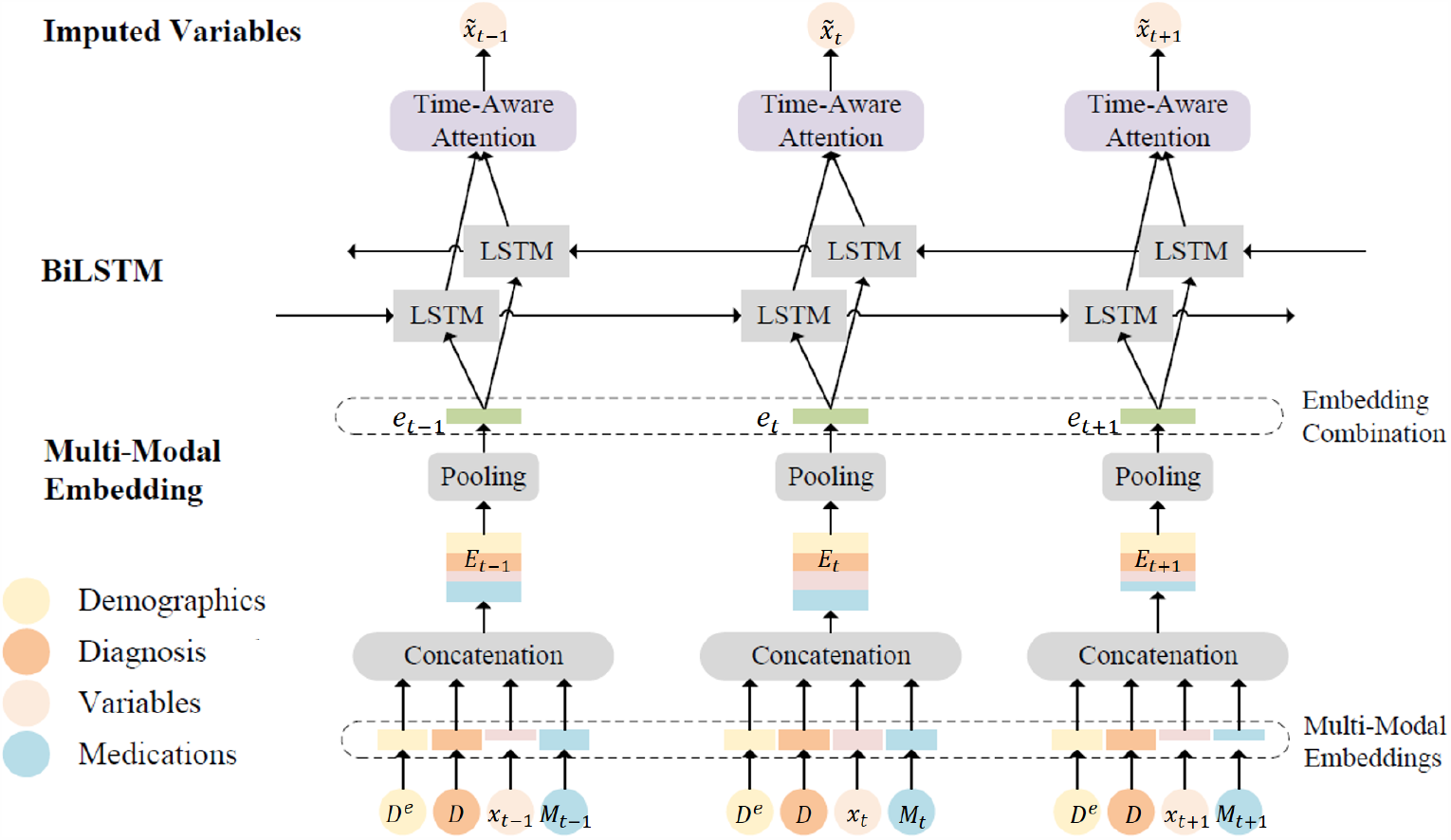
Framework of TAME. TAME takes multi-modal data as inputs, including demographics, diagnoses, variables (i.e., lab tests and vital signs), medications. A patient may take varying numbers of medications and have varying numbers of missing values at different time. Thus, the input dimensions can vary across collections. By concatenating the embeddings of the inputs, we obtain a matrix *E*_*t*_ containing multi-modal information. A pooling layer is followed to output a fixed-size vector *e*_*t*_, which is sent to BiLSTM. At last, a time-aware attention module is used to attend the longitudinal information and then impute the missing values.

#### 2.1.1 Basic Notations

A patient has demographics information *D*^*e*^ (including age *a* and gender *g*) and at least one admission. Following [14] and [8], our subtyping framework treats various admissions of the same patients as different samples. In an admission, the patient has diagnoses *D* = [*d*_1_, *d*_2_, …, *d*_|*D*|_] ∈ *N*^|*D* |^, a collection of variables (i.e., lab test data and vital sign data), denoted by 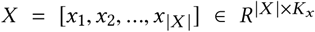, and medications *M* = [*m*_1_, *m*_2_, …, *m*_| *M*|_] ∈ *N*^|*M* |^. At time *t*, the patient is taking a set of medications *M*_*t*_ ⊆ {1, 2, …, |*M*|}.

To address the missing values, we introduce two masking matrix 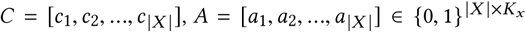 to indicate whether the values in *X* are missing or not. It is initialized as follows:

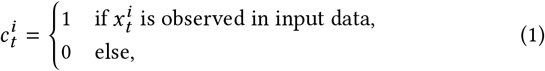

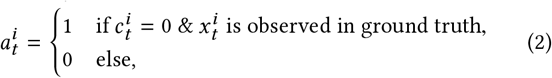

where *i* denotes the *i*^*th*^ variable. *A* is only used to compute imputation loss and validate imputation performance.

The time gaps between collections with observed data carries essential information. Hence, we further introduce three time gap vector and matrices 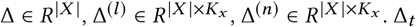 means the time gap between current collection *t* and the last collection 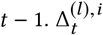 denotes the time gap between current collection *t* and the collection where the *i*^*th*^ variable is observed last time. 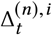 denotes the time gap between current collection *t* and the collection where the *i*^*th*^ variable is observed next time. Δ^(*l*)^ andΔ^(*n*)^ are initialized as follows:

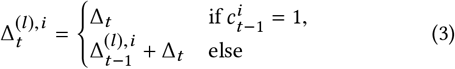

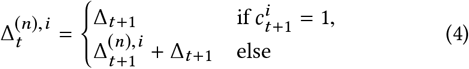

TAME also takes the neighbouring observed values as inputs to incorporate longitudinal information. Thus, we introduce two neighbouring value matrices 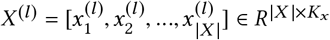 and 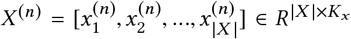, which denote the values observed values of the last and next time.

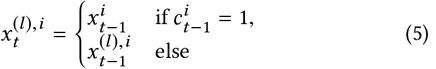

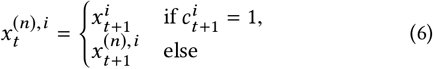

where 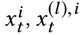 and 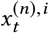 denote the values of the *i*^*th*^ variable of 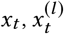 and 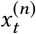 respectively.

#### 2.1.2. Multi-Modal Embedding

We embed multi-modal inputs as vectors and then map them into a same space. A max-pooling layer is followed to combine the multi-modal information.

##### Demographics, Diagnosis and Medication Embedding

For patients’ demographics, their ages are coded to several age groups (i.e., < 30, 30-40, 40-50, etc.). Each patient’s age group and gender are sent to an embedding layer and represented by an embedding matrix 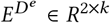. In the same way, we obtain the embeddings of diagnoses, 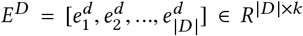, and medications 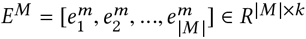.

For medications, we consider their prescription periods but ignore their doses, while diagnoses are valid in the whole admission. At time *t*, the patient is taking a set of medications *M*_*t*_. The corresponding embedding matrix is 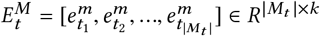, where *t*_*_ ∈ *M*_*t*_

##### Variable Value Embedding

For variables, we propose a novel value embedding to map the values into vectors. Given a variable *i* and the observed values in the whole dataset, we sort the values and discretize the values into *V* sub-ranges with equal number of observed values in each sub-range. The variable *i* is embedded into a vector *e*^*i*^ ∈ *R*^*k*^ with an embedding layer. As for the sub-range *υ*(1 ≤ *υ* ≤ *V*), we embed it into a vector *e*′^*υ*^ ∈ *R*^2*k*^ :

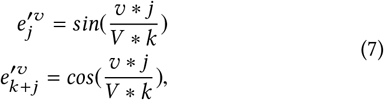

where 0 ≤ *j* < *k*. By concatenating *e*^*i*^ and *e*′^*υ*^, we obtain vector containing both the variable’s and its value’s information. A fully connected layer is followed to map the concatenation vector into a new value embedding vector *e*^*iυ*^ ∈ *R*^*k*^.

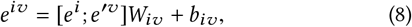

where*W*_*iυ*_ *R*^3*k* ×*k*^, *b*_*iυ*_ *R*^*k*^ are learnable parameters. By stacking the observed values’ embedding vectors in the same collection *t*, we obtain the embedding matrix of the *t*^*th*^ collection variables *x*^*t*^ as 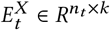, where 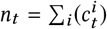. Due to the missing values, the length of 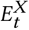 can vary. In the same way, we generate the embeddings of 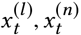 as 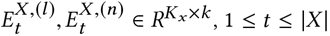.

##### Time Embedding

In order to incorporate the elapsed time between observed values, we present a time embedding for the time gap matrices Δ, Δ^(*l*)^ and Δ^(*n*)^. Given a time gap *δ*, our time embedding layer outputs a vector *e*^*δ*^ ∈ *R*^2*k*^ :

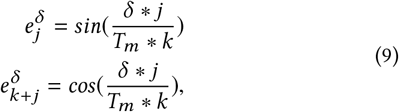

where 0 ≤ *j* < *k, T*_*m*_ denotes the maximum of time gap (0 < *δ*≤ *T*_*m*_). By mapping each time gap value into a vector, we obtain the embeddings of Δ, Δ^(*l*)^, Δ^(*n*)^ as 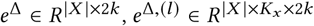 and 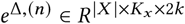.

The proposed value embedding and time embedding remain an important feature of values (time gaps) that similar values (time gaps) are embedded into similar vectors. Moreover, after mapping values into vectors, the embedding bridges multi-modal inputs and makes it possible to handle varying numbers of missing values in different collections.

#### 2.1.3. Multi-modal Embedding Combination

Given the embedding matrices of various EHR data, 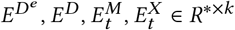, we adopt fully connected layers to project them into a same semantic space. Then we concatenate the results in the new semantic space and obtain a matrix *E*_*t*_ ∈ *R*^*×*k*^, which contains multi-modal information at time *t*. Due to the missing values and varying numbers of medication at different time, the lengths of *E*_*t*_ are varying. A max-pooling layer is followed to map *E*_*t*_ to an vector *e*_*t*_ ∈ *R*^*k*^. We can assume that a well-trained model can ensure that *e*_*t*_ keeps essential information of *E*_*t*_. Then *e*_*t*_ is sent to the LSTM auto-encoder.

#### 2.1.4 BiLSTM Architecture

Given a sequence of multi-modal embedding vectors *e*_*t*_, we build our model based on bidirectional LSTM for its ability to recall long term information. To incorporate the irregular time gaps between successive collections, the time gap embedding vector 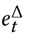 is also input to our auto-encoder. The bidirectional LSTM model can be described as follows:

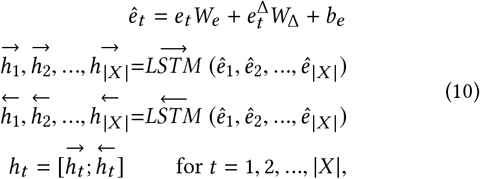

where 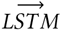 and 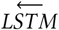 are forward and backward directional LSTM respectively, *W*_*e*_ ∈ *R*^*k* ×*k*^, *W*_Δ_ ∈ *R*^2*k* ×*k*^, *b*_*e*_ ∈ *R*^*k*^ are learnable parameters. *h*_*t*_ ∈ *R*^2*k*^ is the concatenation of 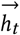 and 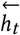.

#### 2.1.5 Time-Aware Attention

In order to incorporate the longitudinal information of observed values, we introduce a time-aware attention module to attend the latest observed values of the variables. Given the time gap embedding matrices 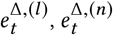, and value embedding matrices 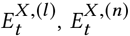, we map the latest observed variables along with their corresponding time gaps into a new space.

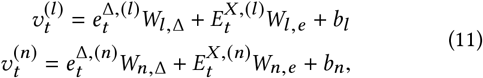

where *W*_*l*,Δ_,*W*_*n*,Δ_ ∈ *R*^2*k* ×*k*^, *W*_*l, e*_, *W*_*n, e*_ ∈ *R*^*k* ×*k*^, *b*_*l*_, *b*_*n*_ ∈ *R*^*k*^ are learnable parameters. By concatenating 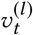 and 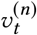, we obtain a embedding matrix 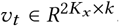. Then the attention mechanism is designed to automatically focus on useful longitudinal information. It takes *h*_*t*_, *υ*_*t*_ as inputs and generate an attention result 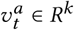.

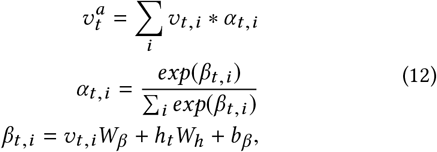

where *W*_*h*_ ∈ *R*^2*k*^, *W*_*β*_ ∈ *R*^*k*^, *b*_*β*_ ∈ *R* are learnable parameters.

#### 2.1.6 Output and Objective Function

Given the LSTM output vector *h*_*t*_ and time-aware attention result 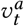, we leverage a fully connected layer to output the missing values.

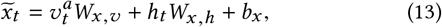

where 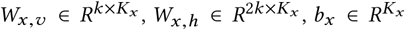 are learnable parameters. The imputation loss is the mean square error between the ground truth 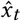 and predictions 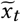 for the *t*^*th*^ collection.

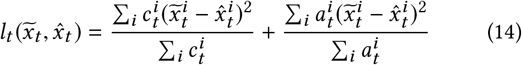

The mean loss of collections is used to train the model. Algorithm 1 in Supplementary Section describes the training process of TAME.

### 2.2 Temporal Similarity with DTW

As is shown in Figure 3, after EHR data imputation, we adopt DTW [13] to compute patient similarity matrix and then cluster the patients with weighted k-means. We replace the missing values with the imputed values and obtain an imputed matrix *S* for each patient.

**Figure 3:**
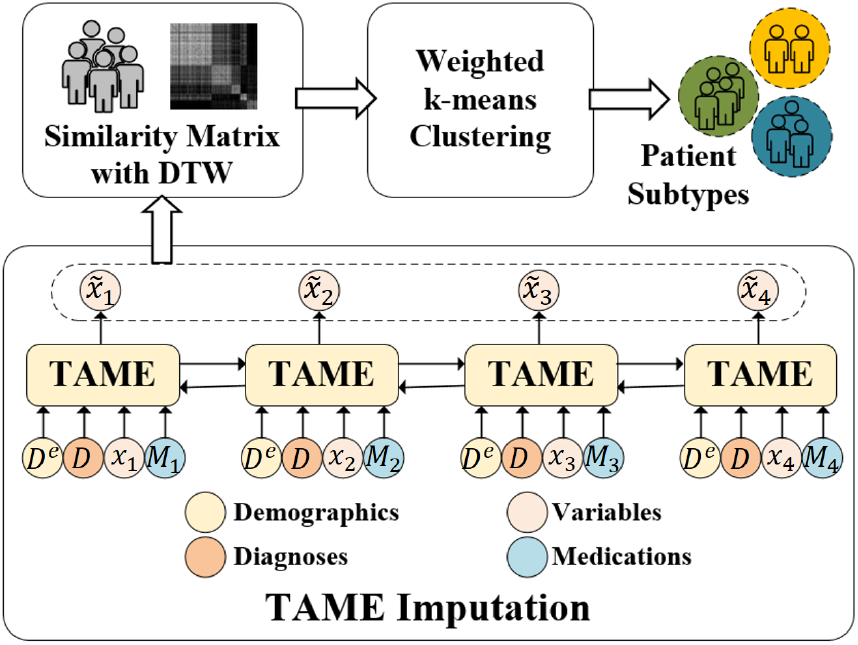
Clustering patients with TAME, DTW and weighed k-means. Taking multi-modal data as inputs, TAME imputes missing values. The imputed results are used to compute temporal similarities between patients with DTW. Weighted k-means is leveraged to cluster the patients into subphenotypes based on the patient similarity matrix.

For patient *i*, his/her imputed matrix is a sequence of variable value vectors, denoted by 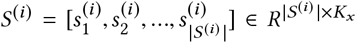. We denote the sub-sequence of 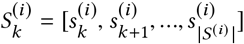. The distance between *S*^(*i*)^ and *S*^(*j*)^ is :

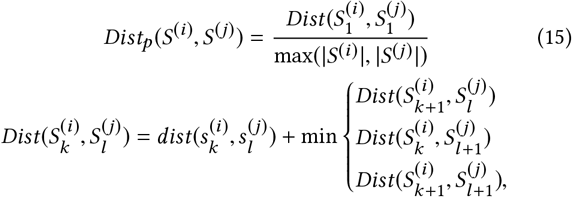

where 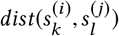 is defined with Euclidean distance:

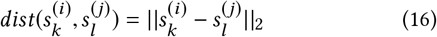

The boundary condition is as follows:

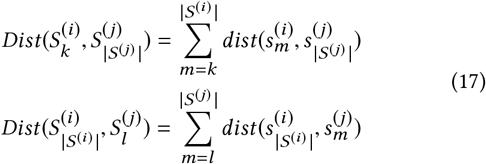

### 2.3 Weighted K-means Clustering

Given the patient similarities (distances), we can leverage k-means to cluster patients into groups. However, the size of patient subphenotypes can be highly imbalanced in clinical settings. For minor groups, outliers are harmful for the calculation of distances between patients and groups. Thus we propose a weighted k-means to mitigate the outliers’ influence by assigning weights for patients in a group when computing the distances. The distance between each patient *S*^(*i*)^ the group *G*_*k*_ is calculated as follows:

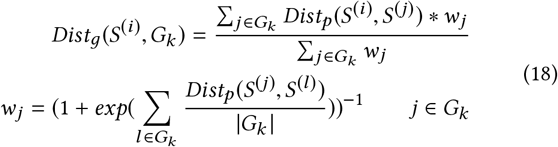

Given the distances between patients and groups, we assign each patient to the group with the smallest distance. Then the distances between patients and new groups are calculated again. The operations repeat until convergence.

## 3 EXPERIMENTS

In this section, we conduct imputation experiments on two EHR datasets, DACMI^1^ and MIMIC-III [9]. Based on the imputed variable values of MIMIC-III data and the computed patient similarity matrix with DTW, we identify subgroups of sepsis patients.

### 3.1 Datasets

Both datasets are publicly available real-world EHRs. The first dataset is DACMI, which contains 13 clinical lab tests that are irregularly measured for 8,267 patients. The statistics of DACMI are listed in Table 4 in Supplementary Section. missing values.

The second dataset is derived from MIMIC-III. We select sepsis patients fulfilling the sepsis-3 criteria [15]. Following [14] and [8], we only focus on adult patients with sepsis. 11,715 sepsis patients are obtained. We use the patients’ data to impute the missing values of 27 sepsis-related variables. The statistics of variables are shown in Table 5 in Supplementary Section. We extract 191 kinds of diagnoses and 498 kinds of medications. The diagnoses and medications that appear less than 100 times are removed. To evaluate imputation method performance, we randomly mask an observed value for each variable in each patient’s data. The masked values are used as ground truth.

### 3.2 Methods for Comparison

To validate the performance of the proposed framework for the imputation task, we implement the following models for comparison.

#### Mean

The mean values of variables are used to impute the

#### KNN

The average values of the top *K* most similar collections are used to impute the missing values.

#### 3DMICE [12]

3DMICE combines MICE [3] and Gaussian Process [7] to impute missing values, which integrates cross-variable and longitudinal information.

#### T-LGBM [19]

T-LGBM builds temporal and cross-variable features as inputs, and adopts LightGBM [10] to impute missing values.

#### BRNN [17]

BRNN prefills the missing values for each variable with the last observed value or mean values of the same variable. Taking as inputs the prefilled data, BRNN adopts a Bidirectional RNN to predict the missing values.

#### CATSI [22]

CATSI consists of two major ingredients: the context-aware recurrent imputation and the cross-variable imputation to capture longitudinal information and cross-variable relations respectively. A fusion layer is used to produce the final imputations.

#### DETROIT [21]

DETROIT builds features based on the observed variables inside five latest collections, and then leverages a network of 8 fully-connected layers to predict missing values.

#### BRITS [5]

BRITS adopts bidirectional RNN to impute missing values. Based on the imputed values, BRITS predicts the values again. The accumulated loss is used to train the model.

#### TAME

Time-Aware Multi-modal auto-Encoder (TAME) is our proposed model to impute the missing values. To evaluate the effectiveness of the proposed operations, including time-aware attention, multi-modal feature combination and variable value embedding, we implement another three variant versions of TAME.

#### TAME^−*T*^

TAME^−*T*^ removes the time-aware attention module when imputing missing values.

#### TAME^−*V*^

TAME^−*V*^ removes the variable value embedding. The method prefills the missing values with mean values and takes the prefilled values as inputs but not the value embeddings.

#### TAME^−*M*^

TAME^−*M*^ just takes the variables as inputs but ignores the other modal data when imputing missing values.

### 3.3 Implement Details

We implement our proposed model with Python 2.7.15 and PyTorch 1.3.0^2^. For training models, we use Adam optimizer with a mini-batch of 64 patients. The multi-modal data are projected into a 512-d space (*k* = 512). We train TAME on 1 GPU (TITAN RTX 6000), with a learning rate of 0.001. We randomly divide the datasets into 10 sets. All the experiment results are averaged from 10-fold cross validation, in which 7 sets are used for training every time, 1 set for validation and 2 sets for test. The validation sets are used to determine the best values of parameters in the training iterations. We use MSELoss as loss function to train models.

We normalize the values of variable *i* as follows:

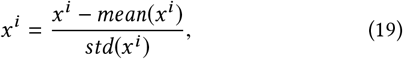

where *mean* and *std* are the mean value and standard deviation for the variable *i* on the whole dataset. When embedding variable values, we discretize the values into 1000 sub-ranges (*V* = 1000) for each variable. We use patients’ first 30 collections data to train TAME and test all collections data for evaluation. For patients with collection length < 30, we pad the data with 0 and set the corresponding values in masking matrices *C* and *A* as 0.

Following [12, 21], we measure the models’ performance with nRMSE. The nRMSE is calculated from the gap between the ground truth and prediction. Given a variable *i*, nRMSE is defined as:

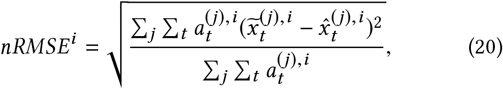

where 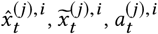 indicate the ground truth, imputed value, and masking indicator for patient *j*, variable *i* in collection *t*. The code and more implementation details are available online^3^.

### 3.4 Results for Imputation

As is shown in Table 1 and Table 2, TAME achieves the best performance for most variables, which demonstrates its effectiveness. Note that due to the space limitation, we display the nRMSE of 14 variables with relatively lower missing rates (50% - 90%) and two mean nRMSE (i.e., the mean nRMSE of 14 variables and 27 variables, denoted by M14 and M27) in Table 2. Due to two variables’ high missing rates (99% for C-Reactive and Bands), M27’s nRMSE is much higher than M14.

**Table 1:** Imputation results for single-modal data on DACMI dataset. The missing rates of these 13 variables are between 1% and 15% as shown in Table 4 in Supplementary Section.

| Method | PCL | PK | PLCO2 | PNA | HCT | HGB | MCV | PLT | WBC | RDW | PBUN | PCRE | PGLU | Mean |
| --- | --- | --- | --- | --- | --- | --- | --- | --- | --- | --- | --- | --- | --- | --- |
| Mean | 0.295 | 0.277 | 0.301 | 0.293 | 0.287 | 0.292 | 0.309 | 0.319 | 0.299 | 0.318 | 0.313 | 0.306 | 0.282 | 0.299 |
| KNN | 0.220 | 0.249 | 0.241 | 0.228 | 0.220 | 0.221 | 0.267 | 0.248 | 0.251 | 0.253 | 0.238 | 0.244 | 0.264 | 0.242 |
| 3DMICE [12] | 0.200 | 0.263 | 0.231 | 0.214 | 0.150 | 0.149 | 0.229 | 0.256 | 0.246 | 0.185 | 0.234 | 0.277 | 0.224 | 0.220 |
| T-LGBM [19] | 0.135 | 0.226 | 0.179 | 0.156 | 0.100 | 0.092 | 0.229 | 0.158 | 0.199 | 0.202 | 0.134 | 0.183 | 0.240 | 0.172 |
| BRNN [17] | 0.155 | 0.230 | 0.196 | 0.174 | 0.090 | 0.087 | 0.245 | 0.175 | 0.211 | 0.208 | 0.154 | 0.210 | 0.252 | 0.184 |
| CATSI [22] | 0.174 | 0.243 | 0.203 | 0.196 | 0.144 | 0.135 | 0.253 | 0.186 | 0.227 | 0.213 | 0.157 | 0.206 | 0.260 | 0.200 |
| DETROIT [21] | 0.138 | 0.219 | 0.172 | 0.155 | 0.093 | 0.087 | 0.234 | 0.152 | 0.199 | 0.201 | 0.137 | **0.181** | 0.262 | 0.172 |
| BRITS [5] | 0.142 | 0.208 | 0.176 | 0.154 | 0.121 | 0.115 | 0.244 | 0.165 | 0.206 | 0.218 | 0.167 | 0.192 | 0.268 | 0.183 |
| TAME<sup>-T</sup> | 0.102 | 0.187 | **0.144** | 0.128 | 0.079 | **0.074** | 0.227 | 0.144 | 0.199 | 0.211 | 0.128 | 0.201 | 0.228 | 0.158 |
| TAME<sup>-V</sup> | 0.121 | 0.193 | 0.164 | 0.145 | 0.081 | 0.076 | 0.238 | 0.168 | 0.203 | 0.201 | 0.141 | 0.210 | 0.222 | 0.166 |
| TAME | **0.100** | **0.179** | 0.155 | **0.125** | **0.073** | 0.077 | **0.218** | **0.136** | **0.198** | **0.180** | **0.121** | 0.185 | **0.221** | **0.151** |

**Table 2:** Imputation results for multi-modal data on MIMIC-III dataset. We impute 27 variables listed in Table 5 in Supplementary Section. Here, we show the nRMSE of 14 variables with relatively lower missing rates (50% - 90%). M14 and M27 denote the mean nRMSE of the 14 and all 27 variables respectively. The full experiments results on 27 variables are available here^3^.

| Method | AG | BCB | CRT | CLR | GLC | HMG | LCT | PLT | PTT | INR | PT | SDM | BUN | WBC | M14 | M27 |
| --- | --- | --- | --- | --- | --- | --- | --- | --- | --- | --- | --- | --- | --- | --- | --- | --- |
| Mean | 0.29 | 0.24 | 0.25 | 0.22 | 0.30 | 0.27 | 0.42 | 0.27 | 0.46 | 0.32 | 0.41 | 0.24 | 0.24 | 0.26 | 0.30 | 0.37 |
| KNN | 0.28 | 0.22 | 0.22 | 0.22 | 0.30 | 0.25 | 0.44 | 0.26 | 0.38 | 0.31 | 0.29 | 0.24 | 0.23 | 0.25 | 0.28 | 0.34 |
| 3DMICE[12] | 0.22 | 0.19 | 0.22 | 0.18 | 0.27 | 0.18 | 0.42 | 0.25 | 0.40 | 0.25 | 0.29 | 0.20 | 0.22 | 0.25 | 0.25 | 0.32 |
| BRNN [17] | 0.15 | 0.17 | 0.20 | 0.13 | 0.29 | 0.12 | 0.40 | 0.20 | 0.41 | 0.16 | 0.24 | 0.18 | 0.17 | 0.26 | 0.22 | 0.30 |
| CATSI [22] | 0.12 | 0.12 | 0.22 | 0.13 | 0.29 | 0.14 | 0.41 | 0.22 | 0.42 | 0.20 | 0.25 | 0.18 | 0.20 | 0.23 | 0.22 | 0.29 |
| DETROIT [21] | **0.11** | 0.09 | 0.28 | 0.09 | 0.27 | 0.13 | 0.38 | 0.22 | 0.46 | 0.17 | 0.24 | **0.10** | 0.17 | 0.22 | 0.21 | 0.27 |
| BRITS [5] | 0.12 | **0.08** | 0.23 | 0.12 | 0.27 | 0.12 | 0.39 | 0.20 | 0.41 | 0.18 | 0.24 | 0.16 | 0.20 | 0.20 | 0.21 | 0.28 |
| TAME<sup>-T</sup> | 0.13 | 0.11 | 0.24 | 0.10 | 0.25 | 0.11 | 0.34 | 0.19 | **0.36** | 0.21 | 0.24 | 0.11 | 0.17 | 0.20 | 0.20 | 0.26 |
| TAME<sup>-V</sup> | 0.16 | 0.13 | 0.23 | 0.12 | 0.26 | 0.11 | 0.36 | 0.20 | 0.38 | 0.19 | 0.22 | 0.14 | 0.17 | 0.20 | 0.21 | 0.26 |
| TAME<sup>-M</sup> | 0.12 | 0.10 | 0.21 | **0.08** | **0.24** | 0.11 | **0.34** | 0.19 | 0.37 | 0.18 | 0.23 | 0.12 | 0.17 | **0.19** | 0.19 | 0.27 |
| TAME | 0.11 | 0.09 | **0.19** | 0.08 | 0.26 | **0.09** | 0.35 | **0.18** | 0.38 | **0.15** | **0.20** | 0.10 | **0.14** | 0.21 | **0.18** | **0.25** |

The overall performance of traditional machine-learning approaches is worse than the deep learning approaches. Mean and KNN do not capture the longitudinal and cross-variable relations, which are essential for accurate imputation. 3DMICE considers the relations, but cannot model the patients’ health state trends like RNN. T-LGBM achieves comparable performance to deep learning models. But the feature engineering of T-LGBM is complicated and hand-designed, which limits its generalization to other datasets. Since we don’t have access to its feature engineering details, the experiment of T-LGBM is not conducted on MIMIC-III dataset. Its results on the DACMI dataset are obtained from [19].

With the consideration of the longitudinal and cross-variable information, the four deep learning baselines perform much better than the traditional machine learning models on average. Among the four deep learning methods, DETROIT directly predicts missing values based on latest collections’ values, which makes it easier to learn longitudinal information. BRITS considers the time gaps between collections, which is also helpful to learn longitudinal information. Thus, the two models outperform the other deep learning baselines. However, all the baselines suffer from the same limitation that their networks take fixed-size vectors as inputs and therefore need to prefill the missing values. BRNN and DETROIT explicitly prefill the missing values with mean or the last observed values. CASTI and BRITS introduce a mask value to indicate whether each variable is observed (= 1) or not (= 0); they take the product of the observed value and mask value as input. This masking operation is the same as to prefill the missing values with 0. Their prefilling operations bring data bias, which limits the models’ performance. Moreover, they do not incorporate other modal data (e.g., diagnoses and medications), which also contain a lot of patients’ health state information and can help impute the missing values.

TAME outperforms the baselines for most variables. TAME can handle varying numbers of missing variables with value embedding without any prefilling operation, which avoids the prefilling bias and thus can improve the imputation performance. By removing the value embedding, there is an obvious performance decline of TAME^−*V*^ compared to TAME. Value embedding also makes it possible to combine multi-modal inputs by mapping all the embeddings into a same space. Note that there isn’t multi-modal data such as diagnoses and medications on the DACMI dataset, and the three versions of TAME only take the single-modal variables data as inputs. By comparing the performance of TAME and TAME^−*M*^ on MIMIC-III dataset, we find that multi-modal inputs can also improve the imputation results. Moreover, our time-aware attention module explicitly attends the latest observed values, which makes it easier to capture the longitudinal information. Thus compared to TAME^−*T*^, TAME achieves better performance.

### 3.5 Sepsis Subtyping

Sepsis subphenotypes are usually identified based on vital signs and lab tests data. Early identification of sepsis subphenotypes is a crucial factor in improving the treatment outcomes. To demonstrate the effectiveness of our early sepsis subtyping framework, we only use patients’ data available up to the first 24 hours in ICUs to impute the missing values and to group patients with the imputed results.

We leverage TAME to impute the missing values and replace them with imputed results. Then, we adopt DTW to compute the patient similarity matrix. Finally, we use a weighted k-means to cluster the sepsis patients into subphenotypes. In this subsection, we conduct experiments to demonstrate whether well-imputed data can help the following subtyping task. Thus, we compare TAME with other strategies to handle missing values: **No-Imp** (we ignore the variables with missing values when computing patient similarity, and do not impute the values) and **Mean-Imp** (we impute the missing values with mean values of variables). Moreover, we also compare weighted k-means (wk-means) with the traditional k-means method. Both our framework and baselines adopt DTW to compute temporal similarity for longitudinal EHR data of patients.

K-means based models need a suitable *K* value when conducting clustering experiments. We group patients into different *K* clusters and then compute the average *P*− *value* for the clustering results. As is shown in Figure 6 in Supplementary Section, *K* = 4 is the best option. Given the imputed data and *K*, our clustering results are shown in Figure 4. Because there is no label for the patient subtyping task, we cannot measure the models’ performance with metrics like Rand Index or NMI which require the knowledge of the ground truth classes. We evaluate our framework and the baselines with two popular metrics Calinski-Harabasz Index (CHI) [4] and Davis-Bouldin Index (DBI) [6], which can measure the performance of clustering algorithms on label-unknown dataset. Note that CHI is related to the size of the dataset, we normalize the value by dividing CHI by the number of the patients. As is shown in Figure 4, **TAME & wk-means** performs the best, which demonstrates the proposed framework’s effectiveness. TAME based frameworks achieve better performance than other imputation based frameworks, which demonstrates that well-imputed data do improve the patient subtyping outcomes. Moreover, by comparing to the results of k-means based frameworks, the weighted k-means based frameworks’ results show better performance, which demonstrates the effectiveness of the weighted k-means in the sepsis subtyping task.

**Figure 4:**
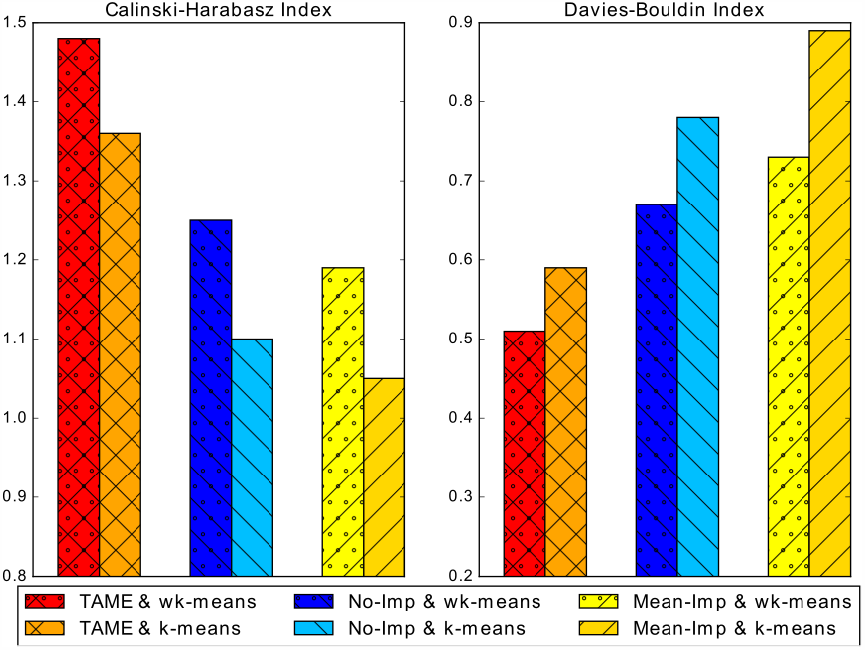
Clustering results on Calinski-Harabasz Index and Davis-Bouldin Index. Note that higher Calinski-Harabasz Index and lower Davies-Bouldin Index relate to a model with better separation between the clusters.

#### 3.5.1 Subphenotypes Analysis

After subtyping the septic patients, we further analyze the subtyping results. All variables used in the clustering method show significant difference across clusters (all *P*− *values* < 0.01 and average *P*− *value* = 4.1 ×10^−5^ as shown in Figure 6 (*K* = 4) in Supplementary Section). The proposed subtyping framework uncovers four subphenotypes with distinct organ dysfunction patterns in septic patients. Following [18], we calculate the Sepsis-related Organ Failure Assessment (SOFA) scores, which are used to describe patients’ organ dysfunction, for the four subphenotypes and overall sepsis population. The details of SOFA computation can be found in Table 6 in Supplementary Section. The characteristics of the subphenotypes are given in Table 3. The subphenotypes have been found to represent the following:

**Table 3:** Cluster descriptive statistics of sepsis subphenotypes. We group patients into four subphenotypes based on their first 24 hours data in ICUs, and display the average of maximum SOFA scores (including six SOFA components and total SOFA) during the whole ICU stays (range from 1 to 153 days) of patients in each subphenotype and overall sepsis population.

|  | Minimal Organ<br>Dysfunction | Renal<br>Dysfunction | CNS & Respiration<br>Dysfunction | Maximal Organ<br>Dysfunction | Overall Sepsis<br>Population |
| --- | --- | --- | --- | --- | --- |
| AVG Age | 59.5 | 62.4 | 59.2 | 62.5 | 60.7 |
| Number of Patients | 4,342 | 3,164 | 2,410 | 1,799 | 11,715 |
| Male/Female | 64%/36% | 54%/46% | 57%/43% | 60%/40% | 59%/41% |
| ICU Mortality Rate | 4.1% | 8.7% | 15.9% | 42.4% | 13.6% |
| Respiration SOFA | 1.5 | 1.6 | **2.3** | **2.3** | 1.8 |
| Coagulation SOFA | 0.6 | 0.8 | 0.9 | **1.6** | 0.9 |
| Liver SOFA | 0.3 | 0.4 | 0.6 | **1.5** | 0.5 |
| Cardiovascular SOFA | 1.4 | 2.0 | 1.9 | **3.0** | 1.9 |
| CNS SOFA | 2.7 | 2.7 | **3.6** | **3.2** | 2.9 |
| Renal SOFA | 3.1 | **3.5** | 3.1 | **3.8** | 3.3 |
| Total SOFA | 8.7 | 10.1 | **11.0** | **14.1** | 10.4 |

**Table 4:** Statistics of Variables in DACMI.

|  | Min | 25%-75% | Max | Missing Rate |
| --- | --- | --- | --- | --- |
| PCL | 62 | 100-108 | 151 | 1% |
| PK | 1 | 3.70-4.40 | 13.2 | 1% |
| PLCO2 | 5 | 22-28 | 65 | 1% |
| PNA | 96 | 135-142 | 179 | 1% |
| HCT | 8.4 | 26.8-32.7 | 77.7 | 13% |
| HGB | 0 | 8.90-11 | 20.8 | 15% |
| MCV | 0 | 86-94 | 139 | 15% |
| PLT | 5 | 130-330 | 2001 | 15% |
| WBC | 0 | 7.1-14.1 | 325.7 | 15% |
| RDW | 0 | 14.5-17.4 | 35.1 | 15% |
| PBUN | 1 | 16-43 | 271 | 1% |
| PCRE | 0 | 0.70-1.90 | 138 | 1% |
| PGLU | 4 | 100-148 | 3565 | 3% |

**Table 5:** Statistics of extracted variables and demographics used to subtype sepsis patients.

|  | Min | 25%-75% | Max | Missing Rate |
| --- | --- | --- | --- | --- |
| Gender | - | - | - | 0 |
| Age | 18 | 53-77 | 89 | 0 |
| Heart Rate | 0.35 | 75-98 | 285 | 21% |
| Respratory | 0.17 | 16-24 | 69 | 21% |
| Temperature | 15 | 36.5-37.6 | 42.2 | 28% |
| WBC | 0.10 | 7.40-14.2 | 471.7 | 69% |
| Bands | 0.80 | 2-11 | 79 | 99% |
| C-Reactive | 0.10 | 15.7-122.8 | 299 | 99% |
| BUN | 1 | 15-40 | 290 | 66% |
| MeanBP | 0.20 | 67-88 | 299 | 26% |
| GCS | 3 | 8-15 | 15 | 33% |
| Urineoutput | 0 | 43-160 | 1200 | 33% |
| Creatinine (CRT) | 0.05 | 0.70-1.80 | 138 | 80% |
| Platelet (PLT) | 5 | 120-287 | 2292 | 82% |
| Glucose (GLC) | 38.0 | 105-157 | 578 | 36% |
| Sodium (SDM) | 74 | 136-142 | 184 | 65% |
| Hemoglobin (HMG) | 1.60 | 9-11.20 | 21.6 | 69% |
| Chloride (CLR) | 39 | 100-108 | 155 | 66% |
| Bicarbonate (BCB) | 2 | 22-28 | 65 | 67% |
| Lactate (LCT) | 0.05 | 1.2-2.9 | 36 | 89% |
| INR | 0.10 | 1.20-1.70 | 48.8 | 80% |
| PTT | 0.15 | 28.5-51.8 | 150 | 79% |
| Magnesium | 0 | 1.80-2.20 | 43.5 | 69% |
| Aniongap (AG) | 1 | 11-15 | 77 | 67% |
| Hematocrit (HMT) | 2 | 26.8-32.9 | 67 | 64% |
| PT | 7 | 13.3-17.8 | 150 | 80% |
| SysBP | 0.06 | 105-136 | 340 | 22% |
| DiasBP | 0.41 | 51-69 | 297 | 22% |
| SPO2 | 1 | 96-99 | 100 | 21% |

**Table 6:** The definition of SOFA score and its components across six organ systems. Each SOFA component score ranges from 0 (normal) to 4 (most abnormal). The total SOFA score ranges from 0 (normal) to 24 (most abnormal).

| SOFA score | 1 | 2 | 3 | 4 |
| --- | --- | --- | --- | --- |
| Respiration |  |  |  |  |
| PaO<sub>2</sub>/FiO<sub>2</sub>, mmHg | < 400 | < 300 | < 200 | < 100 |
| Coagulation |  |  |  |  |
| Platelets ×10<sup>3</sup> /mm<sup>3</sup> | < 150 | < 100 | < 50 | < 20 |
| Liver |  |  |  |  |
| Bilirubin, mg/dl<br>(μmol/l) | 1.2 - 1.9<br>(20 - 32) | 2.0 - 5.9<br>(33 - 101) | 6.0 - 11.9<br>(102 - 204) | > 12.0<br>(> 204) |
| Cardiovascular |  |  |  |  |
| Hypotension | MAP < 70 mmHg | Dopamine ≤ 5<br>or dobutamine (any dose) | Dopamine > 5<br>or epinephrine ≤ 0.1<br>or norepinephrine ≤ 0.1 | Dopamine > 15<br>or epinephrine > 0.1<br>or norepinephrine > 0.1 |
| Central nervous system (CNS) |  |  |  |  |
| Glasgow Coma Score (GCS) | 13 - 14 | 10 - 12 | 6 - 9 | <6 |
| Renal |  |  |  |  |
| Creatinine, mg/dl<br>(μmol/l) or urine<br>output | 1.2 - 1.9<br>(110 - 170) | 2.0 - 3.4<br>(171 - 299) | 3.5-4.9<br>(300 - 440)<br>or < 500 ml/day | > 5.0<br>(> 440)<br>or <200 ml/day |

- Minimal Organ Dysfunction: The subphenotype has the most patients, and the lowest SOFA scores and mortality rate.
- Renal Dysfunction: The subphenotype has a higher Renal SOFA score than the average score of the overall septic population.
- CNS & Respiration Dysfunction: The subphenotype has the highest CNS and respiration SOFA scores. The subphenotype’s mortality rate is much higher than the previous two.
- Maximum Organ Dysfunction: The subphenotype has the fewest patients but the highest mortality rate. All SOFA component scores are higher than average SOFA scores.

The four subphenotypes’ ICU mortality rates are shown in Figure 5. The mortality rates vary significantly across the subphenotypes. Minimal Organ Dysfunction and Renal Dysfunction have lower mortality rates (less than 10%), while the other two subphenotypes are related to much higher mortality rates. Most mortality cases suffer mortality in the first two weeks in ICU stays. Especially in the Maximal Organ Dysfunction subphenotype, the mortality rate grows quickly from the first day. Therefore, it is crucial to identify the patients’ subphenotypes in the early stage and assign more precise treatments for them. We further analyze the variables’ distribution across various subphenotypes; the results are shown in Figure 7 in Supplementary Section.

**Figure 5:**
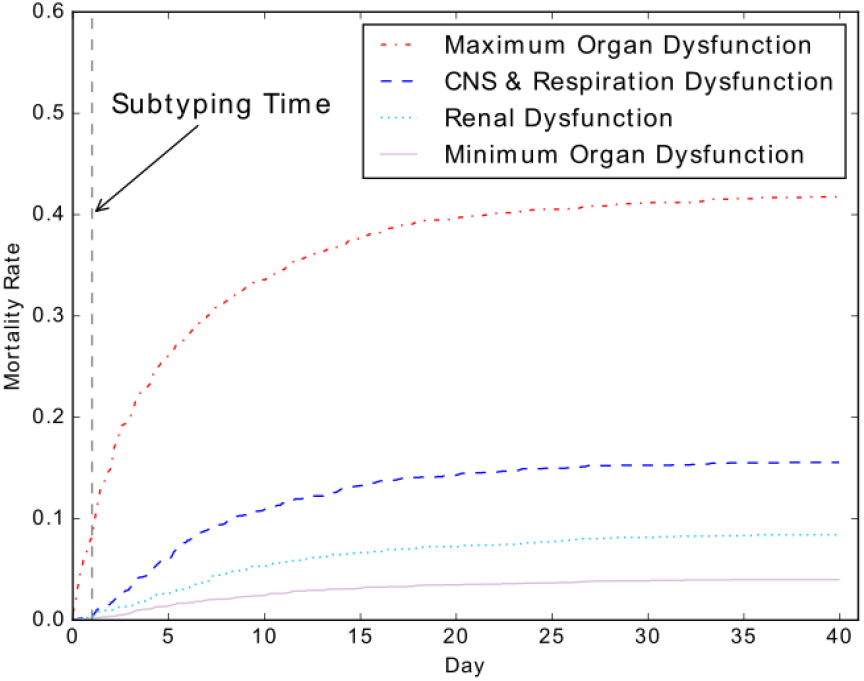
ICU mortality rates of subphenotypes. We only use data available up to the first 24 hours in ICUs for the patient subtyping and show the ICU mortality rates during the whole ICU stays (range from 1 to 153 days). The identified 4 subphenotypes have very different mortality trajectories.

**Figure 6:**
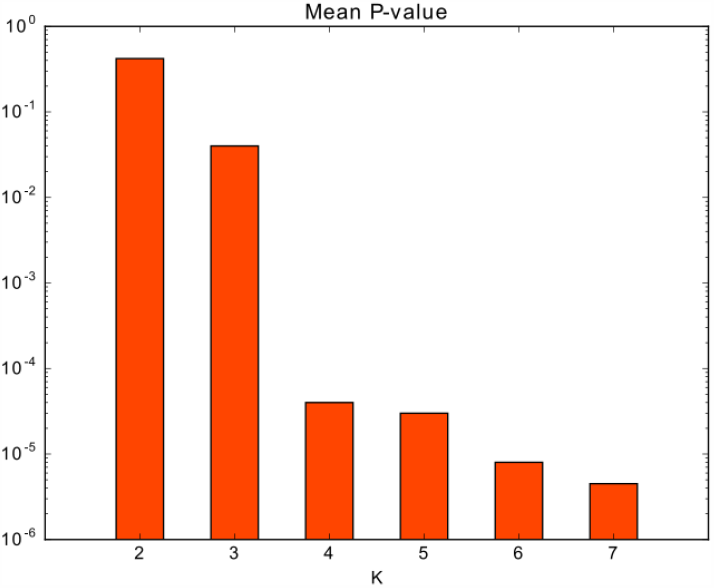
Mean *P*− *value* of variables across different *K* for weighted k-means to cluster the sepsis patients. We select *K* = 4 (which is the elbow point) for the sepsis subtyping task.

**Figure 7:**
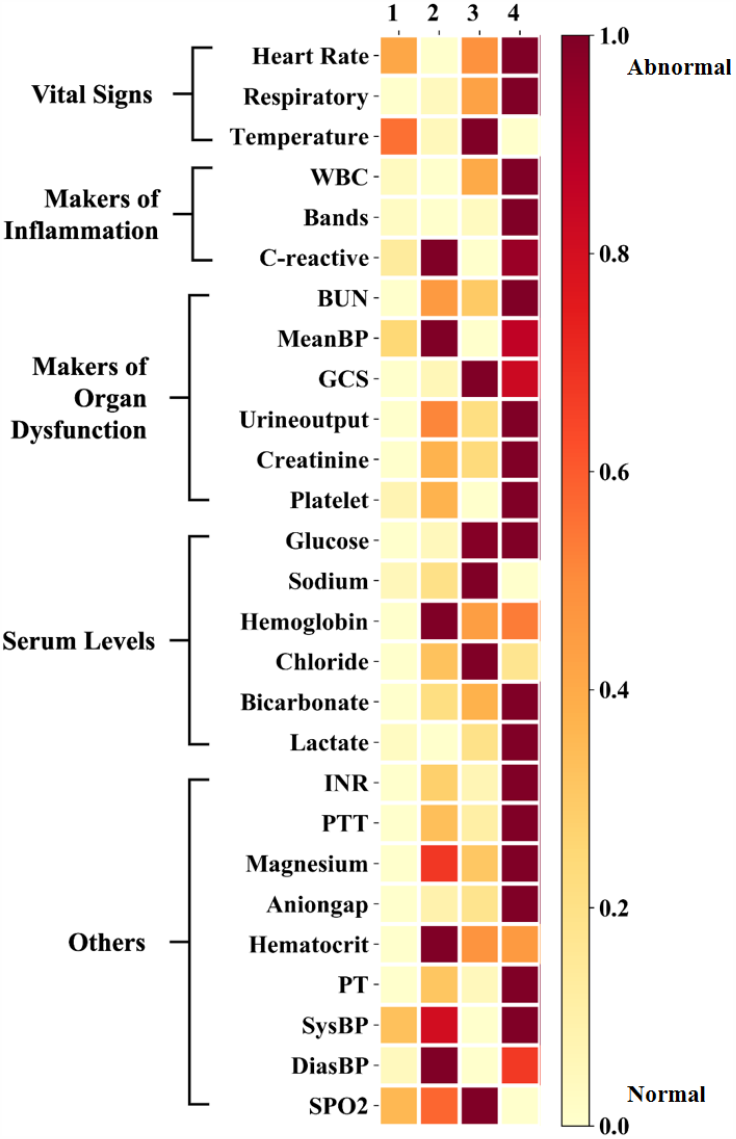
The severity heatmap of the variables across the four subphenotypes. The subphenotypes are 1: Minimal Organ Dysfunction; 2: Renal Dysfunction; 3: CNS & Respiration Dysfunction; 4: Maximum Organ Dysfunction. Deeper color means higher severity.

## 4 RELATED WORK

In this section, we briefly review the existing works related to our models, including patient subtyping and data imputation.

### Patient Subtyping

Identification of sepsis subphenotypes is significant for precise treatments and targeted clinical interventions. During past decades, many studies have focused on patient subtyping with EHR data. Seymour et al. [14] aggregate the values of 29 demographics and variables within the first 6 hours of presentation to the emergency department and adopt k-means to group patients into subphenotypes. Ibrahim et al. [8] cluster patients with a similar method to [14]. The main difference is that Ibrahim et al. [8] use 63 vital sign variable values’ aggregation within the first 24 hours of ICU stays but exclude more patients with high missing rate data. Both papers compute patient similarity based on the aggregation of collections of variables, which ignore the significant temporal information of EHR data. There are also some patient subtyping studies for diseases other than sepsis. Baytas et al. [2] present an auto-encoder model, T-LSTM, to learn a single representation for sequential records of patients, which are then used to cluster patients into clinical subphenotypes. However, T-LSTM only encodes Boolean-value clinical events (e.g., diagnosis codes) but not values of lab tests and vital signs, which are usually used to reflect the health states of patients with sepsis. The model is more suitable for subtype patients with chronic diseases such as heart failure but not acute diseases such as sepsis. In this paper, we propose a new sepsis subtyping framework that computes the temporal similarity between patients and clusters the patients based on the similarity matrix with a weighted k-means.

### Data Imputation

The clinical variables used for similarity measurement and patient subtyping usually have some missing values. Imputation strategies can resolve the problem of missing values in time series data. Early works [1, 16] exploit statistical attributes of observed data, such as mean- and median-filling, which clearly ignore the temporal relations and correlations among variables. 3DMICE [12] combines MICE and Gaussian process to integrate cross-sectional and longitudinal information and achieves better imputation performance. In recent years, deep learning models have become research hotspots and have been applied to time series data imputation. Suo et al. [17] adopt a bidirectional RNN to predict the missing values based on the prefilled data. Yan et al. propose DETROIT [21], which builds features based on the former and latter two collections for each collection. Given the features, DETROIT leverages a network of 8 fully connected layers to predict the missing values. BRITS [5] and CATSI [22] introduce a mask value to indicate whether each variable is observed (= 1) or not (= 0); they take the product of the observed value and mask value as input, then adopt bidirectional recurrent neural networks to model time series data. All the deep learning-based models incorporate longitudinal and cross-variable features. Although the proposed models achieve superior performance in multi-variant imputation tasks, they have two major limitations. The first is that they need to prefill the missing values to provide fixed-size inputs for networks. The second is that they cannot handle multi-modal data (e.g., diagnosis and medications) as inputs, which are probably helpful for imputation in clinical settings. In this work, our proposed TAME well solve the two issues with multi-modal embedding.

## 5 CONCLUSION

We propose a novel clinical data imputation model TAME and a new patient subtyping framework with DTW and weighed k-means. TAME incorporates multi-modal data as inputs, embeds the variables’ values and time gaps, and introduces a time-aware attention mechanism to generate results for missing values. Our model can handle varying numbers of observed variables in different collections without any prefilling operation and capture the cross-variable, cross-modal, longitudinal information. Based on the well-imputed data, we introduce DTW to compute the temporal similarity matrix. Finally, we present a weighted k-means to group sepsis patients with the similarity matrix and identify four meaningful subphenotypes. The subphenotypes show different organ dysfunction patterns and some subphenotypes have a fast-growing mortality rate in the first one or two weeks. The proposed patient subtyping framework is much useful to identify patient subphenotypes in patients’ early stages of ICU stays, which paves the way for improved personalization of sepsis management.

## Data Availability

We used two publicly available datasets: MIMIC-III and DACMI.

https://mimic.physionet.org/

http://www.ieee-ichi.org/2019/challenge.html

## 6 ACKNOWLEDGMENTS

The authors would like to thank Dr. Lawrence Lynn and Ms. Raegan Heitzenrater for the weekly discussions of sepsis and their language editing during the preparation of the manuscript. This project was funded in part under a grant with Lyntek Medical Technologies Inc.

## 7 SUPPLEMENTARY SECTION

### 7.1 TAME Model and Availability

Algorithm 1 describes the overall training process of TAME. The codes will be available at Github. Here is the hyperlink^3^.

#### Algorithm 1

Time-Aware Multi-modal auto-Encoder (TAME)

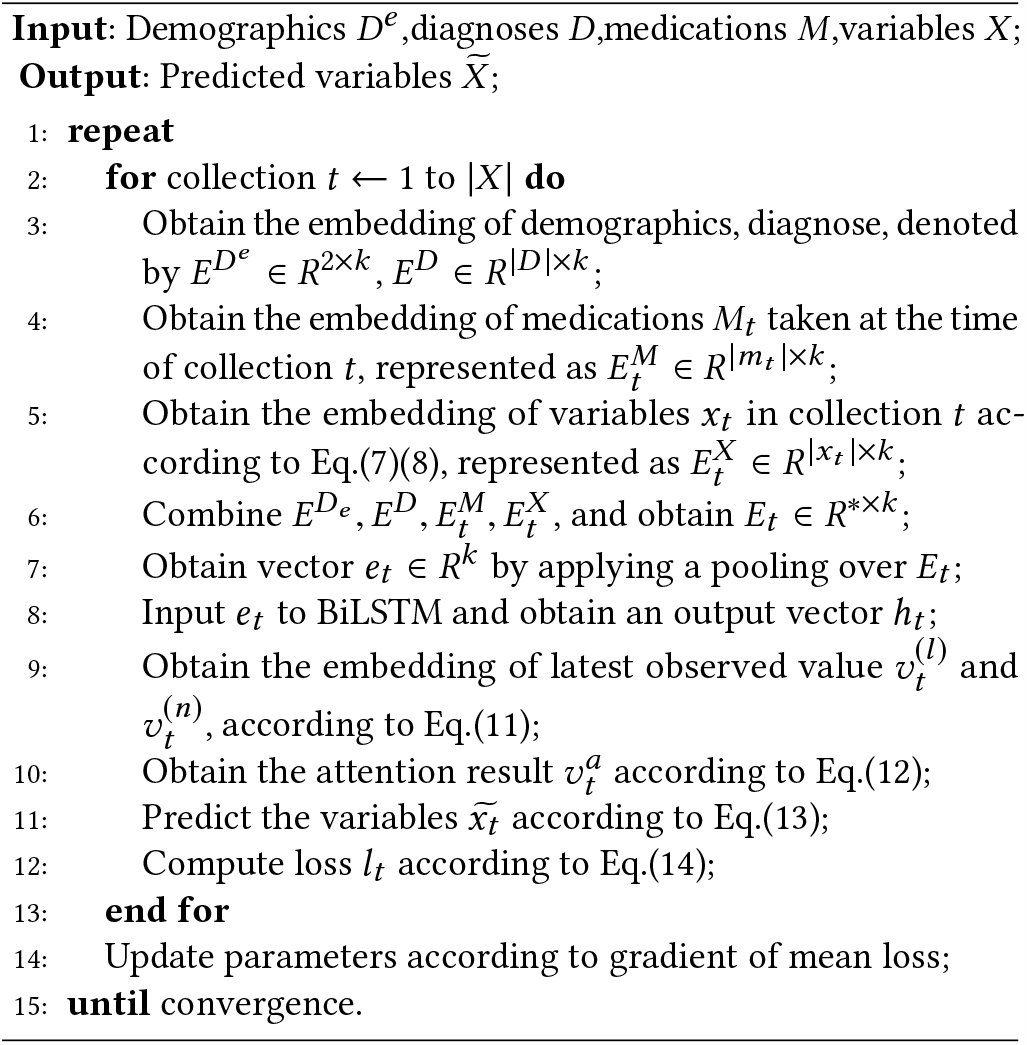

### 7.2 Statistics of DACMI and MIMIC-III Datasets

#### 7.2.1 DACMI Dataset

The 13 variables’ minimums, interquartile ranges, maximums and missing rates on the DACMI dataset are shown in Table 4.

#### 7.2.2 MIMIC-III Dataset

We extract the 2 demographics (i.e., age and gender) and 27 sepsis-related variables (i.e., vital signs and lab tests) of the 11,715 sepsis patients, including 41% female and 59% male. The statistics of extracted demographics and variables are listed in Table 5.

### 7.3 SOFA Score Calculation

We use SOFA (Sepsis-related Organ Failure Assessment) score to describe organ dysfunction in septic patients. Following [18], the six organ SOFA scores are calculated as shown in Table 6. Each organ’s SOFA score ranges from 0 (normal) to 4 (most abnormal). The total SOFA score ranges from 0 (normal) to 24 (most abnormal).

### 7.4 K Value Selection for Weighted K-means

After data imputation and patient similarity computation, we adopt weighted k-means to cluster patients into *K* groups. We conduct experiments with different *K* and compute the mean *P* − *value* for the variables. As is shown in Figure 6, *K* = 4 is the best option. When *K* < 4, the average *P* − *value* becomes much higher (> 0.01). When *K* > 4, there isn’t a large decline on average *P* − *value*.

### 7.5 Sepsis Subphenotype Visualization

After grouping the septic patients into 4 subphenotypes, we compute the average value of variables across subphenotypes. The average severity scores of different variables are shown in Figure 7. Overall the Maximum Organ Dysfunction subphenotype has the most severe states. The CNS & Respiration Dysfunction subphenotype is related to a severe GCS value, which is consistent with its high CNS SOFA component in Table 3. Renal Dysfunction subphenotype has relatively worse Urineouput and Creatinine, which are related to Renal system. The overall variable severity distribution is identical to the SOFA component scores in Table 3.

### Footnotes

1 http://www.ieee-ichi.org/2019/challenge.html

2 https://pytorch.org/

3 https://github.com/yinchangchang/TAME

